# How did the COVID-19 Pandemic impact self-reported cancer screening rates in 12 Midwestern states?

**DOI:** 10.1101/2022.04.13.22273837

**Authors:** Jason Semprini

## Abstract

**Objective:** In the early months of the COVID-19 pandemic, the U.S. healthcare system reallocated resources to emergency response and mitigation. This reallocation impacted essential healthcare services, including cancer screenings.

**Methods:** To examine how the pandemic impacted cancer screenings at the population-level, this study analyzes 2018 and 2020 Behavioral Risk Factor Surveillance System (BRFSS) data to estimate the change in the proportion of eligible adults reporting a recent cancer screen (mammogram, pap smear, colon/sigmoidoscopy, blood stool test). All analyses accounted for response rates and sampling weights, and explored differences by gender and region across 12 Midwestern states.

**Results:** We found that the proportion of adult women completing a mammogram declined across all states (−0.9% to -18.1%). The change in colon/sigmoidoscopies, pap smears, and blood stool tests were mixed, ranging from a 9.7% decline in pap smears to a 7.1% increase in blood stool tests. Declines varied considerably between states and within states by gender or metro/urban/rural status.

**Conclusions:** The COVID-19 pandemic led to delayed breast, cervical, and colorectal cancer detection services. Policymakers should aim to advance cancer control efforts by implementing targeted screening initiatives.

## Background

In March 2020, policymakers, healthcare providers, and public health professionals shifted their resources towards COVID-19 prevention and mitigation. The reallocation was a response to the uncertainty of the pandemic’ s initial projections. ^1-3^ Health systems also began instituting policies to protect at-risk adults from adverse COVID-19 outcomes.^2,4-6^ This required reallocating healthcare capacity, but also limiting exposure to patients and providers by minimizing “non-essential” care.

The U.S. healthcare system’ s response to the COVID-19 pandemic came at the expense of other healthcare services.^7^ Early evidence showed dramatic declines in healthcare service utilization. One report found a thirty-percent reduction in individual-level healthcare consumption.^8^ In March and April 2020, most adults reported having difficulties accessing necessary or choosing to delay care.^9^ Healthcare providers expected these delays to slow population health and equity efforts.^10^

Cancer screenings were among the services delayed due to the pandemic.^10^ However, most studies on cancer services in the early months of the pandemic focused on strategies to maintain safe treatment regimens for cancer patients.^11-14^ As data eventually became available, investigators found that cancer screenings declined substantially.^15^ Bakouny’ s report found that, compared to pre-pandemic months, claims for mammograms, pap smears, and colorectal cancer screenings declined 60-80 percent in March/April 2020. It is unknown how screening rates evolved over the rest of the pandemic’ s first year.^15^

### New Contributions

This study makes many contributions to the evidence on cancer screenings curing the COVID-19 Pandemic. The Bakouny report was conducted in the New England region, with little attention to subgroup analyses.^15^ Should healthcare providers in other U.S. states expect 60-80% declines in cancer screenings? Were the changes in screening rates more pronounced in specific subgroup populations? My study aims to examine how cancer screening rates changed in 12 Midwestern states, stratifying analyses by gender and metro/urban/rural status. The most critical gap I address, however, relates to the data. Existing evidence on healthcare service changes during the pandemic relied on claims data, but how well can we use this evidence to implement interventions that get adults back to screening? Claims data doesn’ t represent the population of adults eligible for screenings, but rather only represents adults accessing the healthcare system. These early claims-based studies can calculate the decline in screenings, but only population-based studies can identify changes in cancer screening rates for adults who delayed or missed care during the pandemic. As we move on from the emergency phase of the pandemic, this study aims to identify which populations’ cancer screening rates were most impacted by the pandemic with the goal of informing targeted return to screening initiatives.

## Materials and Methods

### Data

This study is among the first to analyze the 2020 Behavioral Risk Factor Surveillance System (BRFSS) questionnaire’ s cancer screening module.^16^ BRFSS is a state-based, nationally representative survey implemented by telephone. Each year, BRFSS queries a random sample of adults to represent health risks and behaviors of the U.S. population. On a biannual basis, most states implement a BRFSS questionnaire which asks about cancer screening: mammograms, pap smears, colonoscopies or sigmoidoscopies (colon/sigmoidoscopies), and blood stool tests. All 12 Midwestern states implemented the biannual cancer screening module in 2020.^17^

For each of the four cancer screenings, BRFSS asks eligible adults the duration since their last screening. I used these questions to develop binary measures of annual cancer screening behavior and categorized eligible adults as either having completed a respective cancer screen in the past year, or not.

### Design

The BRFSS survey is implemented in waves and conducted over the course of each year. I leverage the implementation of the survey in my research design. By comparing responses in quarter 1 (Jan-Mar. 2020) with responses in quarter 4 (Oct.-Dec. 2020), I obtain two seemingly comparable groups of adults with different levels of exposure to the COVID-19 pandemic. Adults surveyed in quarter 4 would have endured at least six months of the pandemic whereas the adults surveyed in quarter 1 would have yet to be exposed (note: BRFSS briefly stopped surveying activities after the initial emergency towards the end of March 2020). I account for heterogeneous response rates by first calculating the proportion of all eligible adults reporting a recent cancer screen. Then, I account for seasonal survey implementation heterogeneity by differencing out screening behavior and response rate changes from quarter 1 to quarter 4 in 2018. Finally, to compare how these differences varied relative to pre-pandemic screening rates, I estimate the relative change by dividing the Q4-Q1 difference estimate by the 2020-Q1 baseline screening rate.

Subgroup analyses examine differences by gender (male, female) and region (metro, urban, rural) across each Midwestern state. All calculations utilize BRFSS probability weights to obtain a representative sample of each state’ s population. The analysis was performed in STATA v. 17. For replication purposes, see the dataset creation and analytical code in the appendix. The study used publicly available, deidentified data and is not considered Human Subjects Research or subject to IRB determination.^16^

## Results

### Mammograms

Across all states, the proportion of women completing a recent mammogram declined in 2020. The decline was smallest in Minnesota (−0.9%) and highest in Indiana (−18.1%). These declines represented a 2.4% and 40.0% relative reduction from pre-pandemic baseline screening rates. The changes, however, are mixed when disaggregating by metro, urban, and rural status within these states. The proportion of metro women completing a recent mammogram increased from quarter 1 to quarter 4 in 2020 for two states: Illinois (+0.1%) and South Dakota (+5.2%). Metro mammogram rates declined in all other states, ranging from a 0.3% decline in Michigan to a 16.8% decline in Indiana. The proportion of urban women reporting a recent mammogram increased only in Minnesota (+5.7%), with reductions ranging from 1.2% in Iowa to 22.3% in Wisconsin. The largest changes and greatest variability were observed in rural respondents. The proportion of rural women in Nebraska reporting a recent mammogram increased 16.6%. The proportion of rural women reporting a recent mammogram also increased in Illinois, Iowa, Kansas, Minnesota, and Wisconsin. The proportion declined for rural women across all other states, with the largest decline found in Indiana (−35.9%). See Figure 2 and Supplemental Table 2 for the full set of results.

### Pap Smears

The change in proportion of women reporting a recent pap smear was mixed across states. In Michigan (+7.9%), North Dakota (+1.3%), and Wisconsin (+4.4%) the proportion of women reporting a recent pap smear increased from quarter 1 to quarter 4 in 2020. The proportion of pap smears declined in the other nine states, with declines ranging from 1.0% in Minnesota to 10.0% in South Dakota. These declines represent a 3.2% and 26.1% relative change in pre-pandemic pap smear rates. When examining rates by regional status, there is less volatility in the changes. For example, the highest observed increase in the proportion of women reporting a recent pap smear was 9.9% in metro Kansas, 13.0% in urban Missouri, and 13.5% in rural Missouri. Conversely, the largest declines were found in metro South Dakota (−14.6%), urban Iowa (−10.8%), and rural South Dakota (−12.2%). Figure 2 and Table 2 report the full set of results.

### Colonoscopies & Sigmoidoscopies

Like the estimates for pap smears, the difference in colon/sigmoidoscopies between quarter 1 and quarter 4 varied by states. The proportion of eligible adults reporting a recent colon/sigmoidoscopy increased from quarter 1 to quarter 4 in Kansas (0.3%), Missouri (1.2%) and South Dakota (1.8%). The proportion declined in nine states, with estimates ranging from 0.5% in North Dakota to 8.9% in Minnesota. These declines represent a 1.9% and 31.6% relative reduction the proportion of adults reporting a colon/sigmoidoscopy. The subgroup analyses reveal that the proportion of adults reporting a recent colonoscopy or sigmoidoscopy declined consistently for metro and respondents. Only one state (SD) showed an increase in the proportion of metro colonoscopies and only two states (MO, WI) showed an increase in the proportion of male colonoscopies. There were also only two states reporting increased proportion of colonoscopies for rural respondents (IN, WI). For metro, rural, and male populations, the rest of the states showed declines. For urban and female respondents, however, there was much more heterogeneity across states. See Tables and Figures 3 and 4 for the full set of results.

### Blood Stool Tests

Contrary to the other screenings, most states were found to have increased the proportion of adults reporting a recent blood stool test. These increases ranged from 0.5% in North Dakota and Kansas to 7.1% in Wisconsin. These respective increases represent a 0.7% and 8.9% change relative to pre-pandemic blood stool test rates. The proportion of blood stool tests only declined from quarter 1 to quarter 4 in Indiana (3.0%), Michigan (4.5%), Minnesota (1.4%) and Missouri (3.4%). These changes in blood stool rates varied across states by metro/urban/rural status. More consistent however, were the results disaggregated by gender. The proportion of adult female respondents having completed a recent blood stool test declined in eight states, ranging from a 0.2% reduction in Illinois to a 7.8% reduction in Michigan. Blood stool rates only increased for female respondents in Iowa (+2.7%), Ohio (+3.1%), South Dakota (+3.4%), and Wisconsin (+5.9%). Conversely, the proportion of male respondents reporting a recent blood stool test only declined in Michigan (−1.0%), Minnesota (−0.9%), and Missouri (−3.4%). In the other nine states, the blood stool test rates increased for male respondents, ranging from 1.0% in North Dakota to 9.5% in South Dakota. See figures and tables 3 and 4 for the full set of results.

## Discussion

Policymakers, providers, and public health professionals have already begun to implement return to screening initiatives.^18^ This timely research can inform their ongoing efforts. In summary, the proportion of adult women reporting a recent mammogram declined across all 12 Midwestern states. With a few exceptions, the proportion of adults reporting a recent pap smear declined, as did the proportion of adults reporting a recent colon/sigmoidoscopy. Meanwhile, the rate of blood stool tests generally increased, but by a smaller magnitude than the decline in colon/sigmoidoscopies. We should not expect the rise in blood stool tests to offset the declines in colon/sigmoidoscopy rates.

There appeared to be greater heterogeneity within states (by metro/urban/rural status) for mammograms than for pap smears. However, the clearest pattern which emerged from the data were the changes in colon/sigmoidoscopy rates for metro and rural respondents, and for adult males. There is also stark heterogeneity across states. This research will be most useful for motivating and informing targeted initiatives.

Practitioners and policymakers can use this evidence to identify which screening services declined the most in their state and the population experiencing the greatest declines. Until evidence emerges that screening rates have returned to or exceeded pre-pandemic levels for all groups, stakeholders in these 12 Midwestern states should prioritize the services and populations most impacted by the pandemic.

### Limitations

This study is not without its limitations. First, the experience of adults in 12 Midwestern states may not generalize to other U.S. states or census regions. Moreover, while the Midwestern states provided interesting data for examining differences by metro/urban/rural status, the region is less diverse from a racial/ethnic perspective (at least in the BRFSS data). The small sample size of adults identifying as a race or ethnicity other than non-Hispanic White would yield unstable estimates and unreliable or nonrepresentative sample weights. Future research should investigate how cancer screening rates may have differentially changed by race/ethnic identity. Another limitation stems from the outcome of interest: having complete a cancer screen in the past 12 months, and the timing of survey. Adults in the exposure group (2020-Q4) reporting a recent cancer screen could have completed the screen anytime between October 1^st^, 2019 to December 30^th^, 2020. If the screening was completed in 2019, we may be underestimating the decline in screening rates for 2020. We also have no way to know if these recent cancer screens were completed during the end of 2020 and may therefore be missing a signal of “return to screening” behavior. As new data emerge, future research should continue to survey how screening rates evolve over time. Unfortunately, among all population-based survey data, BRFSS provides the most comprehensive cancer screening data, but the cancer module is only implemented biannually. So, the next BRFSS cancer screening module will not be completed until the end of 2022. The data will not be available to researchers until late 2023.^16^ Despite this study’ s limitations, the reliability of the BRFSS data and simple, yet analytically valid design yield baseline estimates for the pandemic’ s effect on the population’ s screening rates as Americans emerge from a two-year public health emergency.

## Conclusion

The covid-19 pandemic forced adults in all 12 Midwestern states to delay critical cancer screening services. Fewer breast, cervical, and colorectal cancer screenings in 2020 may lead to more cancer diagnoses in the coming years, likely at more advanced and aggressive stages. Policymakers can use evidence from this study to implement targeted screening initiatives and mitigate the pandemic’ s long-term impact on cancer control systems.

## Data Availability

All BRFSS data are publicly available from the CDC. https://www.cdc.gov/brfss/index.html

https://github.com/jsemprini/brfss-covid-screening-midwest

