## Supplementary material for "How did the COVID-19 Pandemic impact self-reported cancer screening rates in 12 Midwestern states?": Figures 1-4

**Figure 1 –Self-Reported Changes in 2020 Cancer Screening Rates**

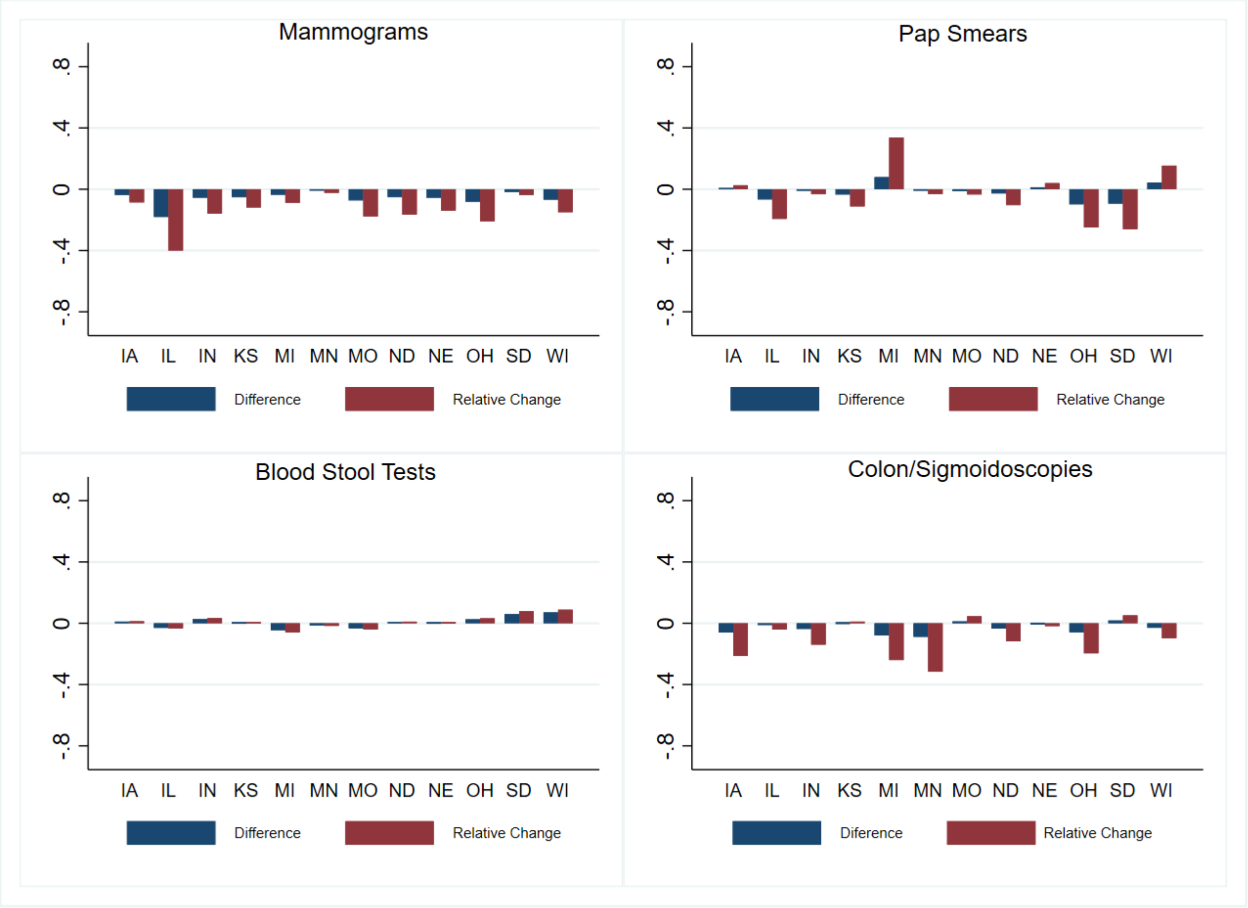


*Figure 1 visually presents the differences between 2020-Q4 and 2020-Q1 self-reported cancer screening rates. Figure 1 also depicts the relative change of this difference as a percentage of 2020-Q1 screening rates.*

**Figure 2 – Self Reported Changes in 2020 Mammograms and Pap Smears – by Metro, Urban, Rural Status**

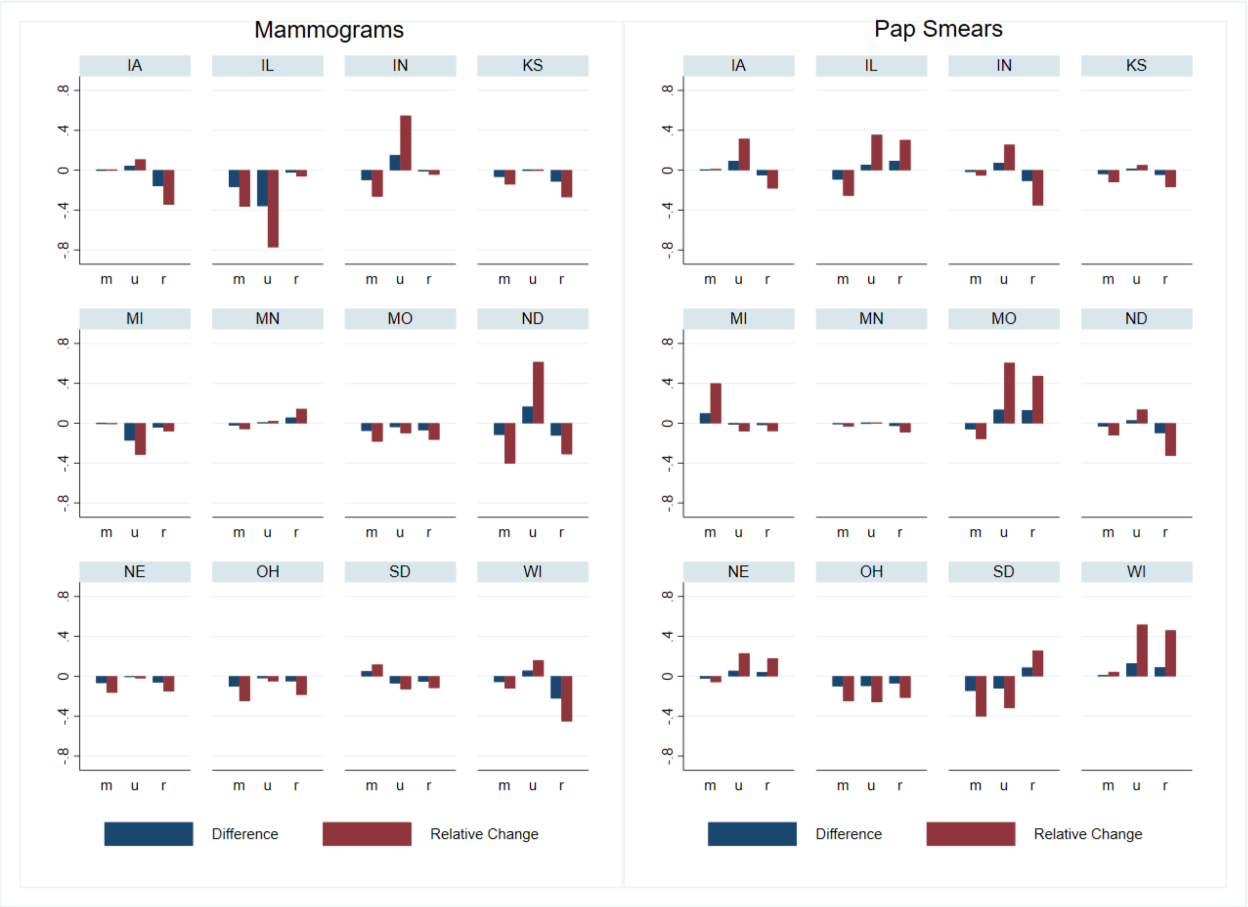


*Figure 2 visually presents the differences between 2020-Q4 and 2020-Q1 self-reported mammogram and pap smear rates. Figure 2 also depicts the relative change of this difference as a percentage of 2020-Q1 screening rates. Sample restricted to adult female respondents. Region status derived from BRFSS data. m = metro, u = urban, r = rural.*

**Figure 3 – Self Reported Changes in 2020 Blood Stool & Colon/Sigmoidoscopies Rates – by Metro, Urban, Rural Status**

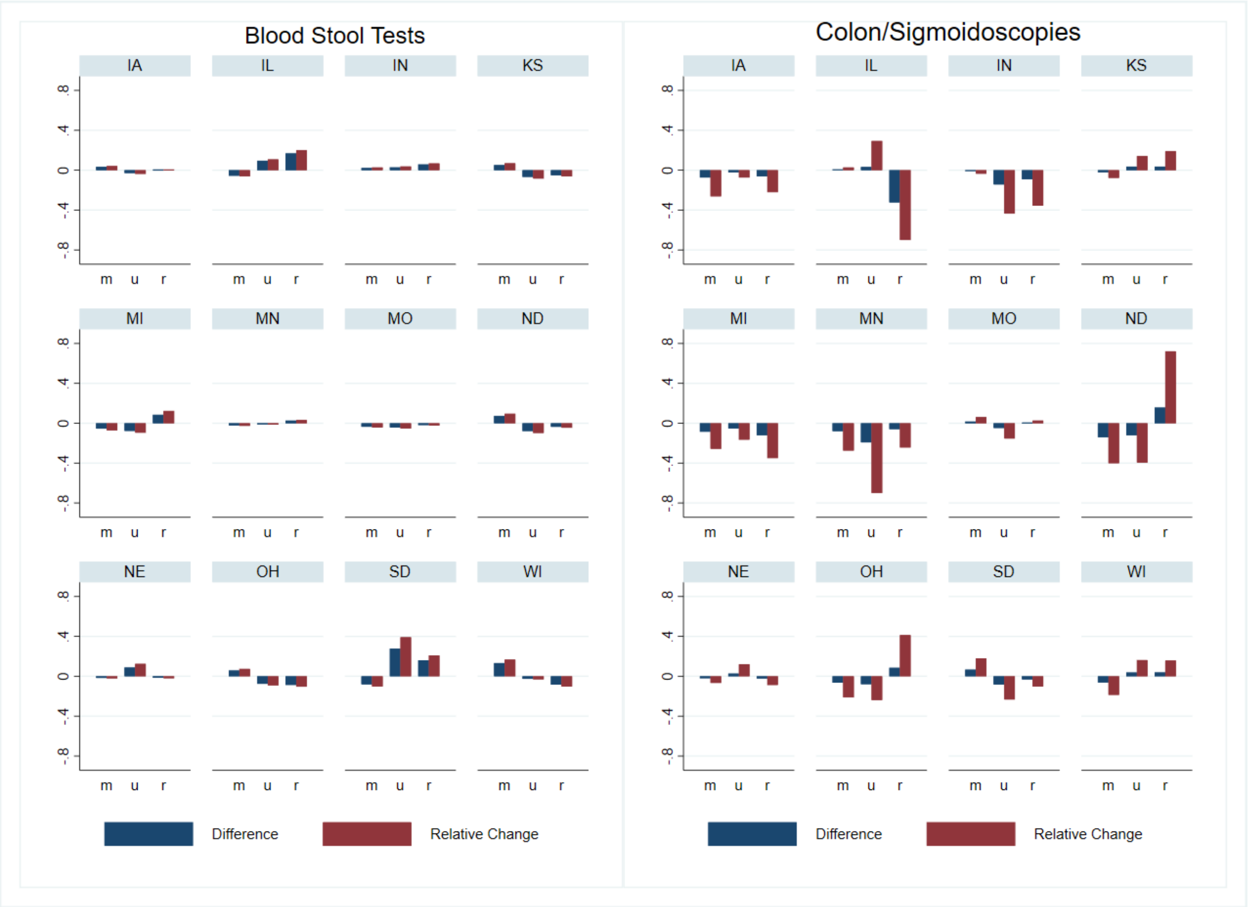


*Figure 3 visually presents the differences between 2020-Q4 and 2020-Q1 self-reported blood stool tests and colon/sigmoidoscopy rates. Figure 3 also depicts the relative change of this difference as a percentage of 2020-Q1 screening rates. Sample includes male and female respondents. Region status derived from BRFSS data. m = metro, u = urban, r = rural.*

**Figure 4 – Self Reported Changes in 2020 Blood Stool Tests & Colon/Sigmoidoscopies – by Sex**

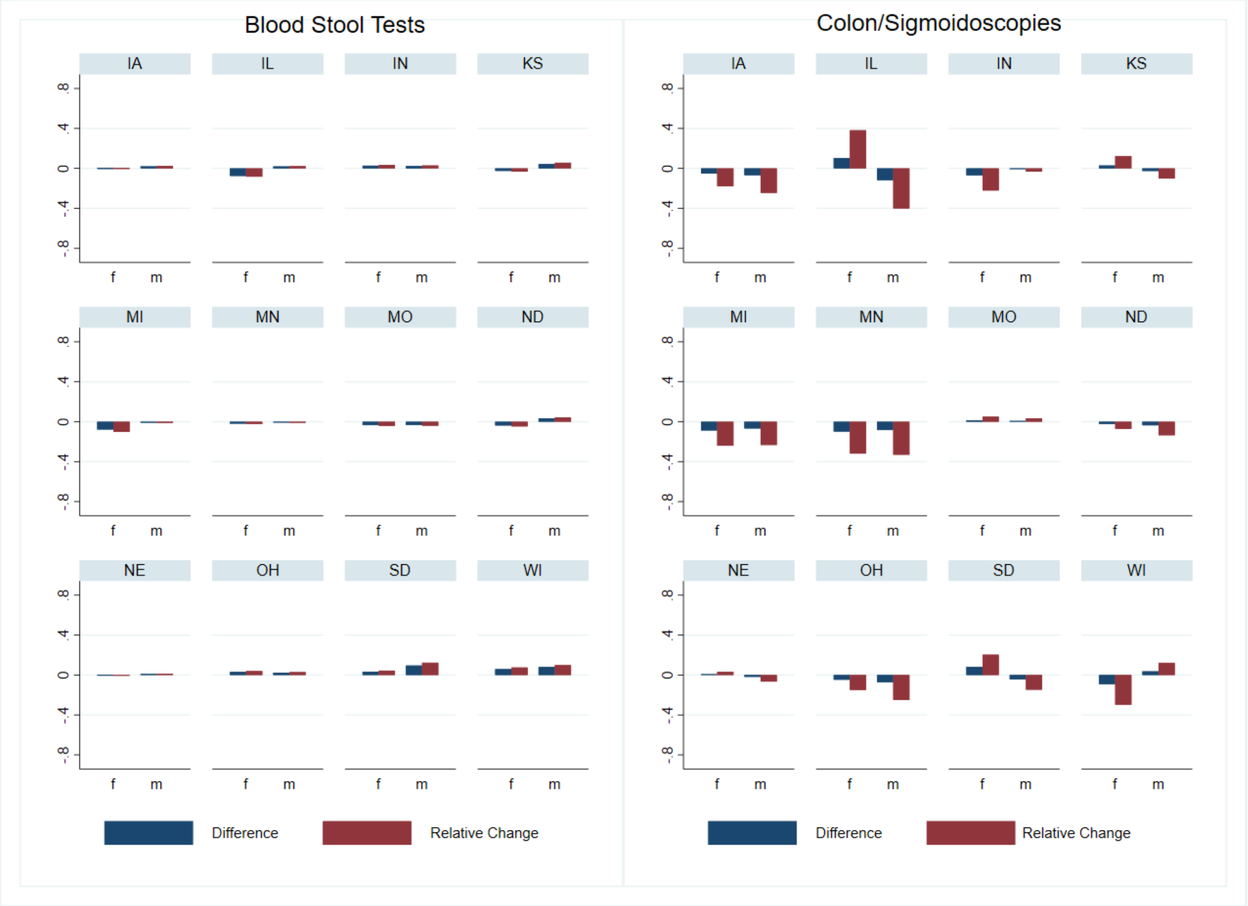


*Figure 4 visually presents the differences between 2020-Q4 and 2020-Q1 self-reported blood stool tests and colon/sigmoidoscopy rates. Figure 4 also depicts the relative change of this difference as a percentage of 2020-Q1 screening rates. Sex determined by self-reported BRFSS data. f = female, m = male.*
