## Supplemental Tables 1-6 for "How did the COVID-19 Pandemic impact self-reported cancer screening rates in 12 Midwestern states?"

**Supplemental Table 1 – Change in Self-Reported Difference in Cancer Screening Rates in 2020 – Full Sample of Eligible Adults**

| | Mammograms | | Pap Smears | | Blood Stool Tests | | Colon/Sigmoidoscopies | |
|---|---|---|---|---|---|---|---|---|
| | Difference | Relative Change | Difference | Relative Change | Difference | Relative Change | Difference | Relative Change |
| *IL* | -0.038 | -0.086 | 0.008 | 0.026 | 0.010 | 0.013 | -0.060 | -0.213 |
| *IN* | -0.181 | -0.400 | -0.067 | -0.194 | -0.030 | -0.034 | -0.012 | -0.041 |
| *IA* | -0.056 | -0.159 | -0.010 | -0.033 | 0.027 | 0.034 | -0.037 | -0.140 |
| *KS* | -0.052 | -0.119 | -0.034 | -0.112 | 0.005 | 0.007 | 0.003 | 0.011 |
| *MI* | -0.036 | -0.089 | 0.079 | 0.337 | -0.045 | -0.060 | -0.079 | -0.240 |
| *MN* | -0.009 | -0.024 | -0.010 | -0.032 | -0.014 | -0.017 | -0.089 | -0.316 |
| *MO* | -0.073 | -0.177 | -0.012 | -0.034 | -0.034 | -0.040 | 0.012 | 0.046 |
| *NE* | -0.052 | -0.166 | -0.027 | -0.104 | 0.008 | 0.010 | -0.034 | -0.118 |
| *ND* | -0.056 | -0.140 | 0.013 | 0.040 | 0.005 | 0.006 | -0.005 | -0.019 |
| *OH* | -0.082 | -0.209 | -0.098 | -0.250 | 0.026 | 0.032 | -0.059 | -0.196 |
| *SD* | -0.018 | -0.039 | -0.094 | -0.261 | 0.061 | 0.080 | 0.018 | 0.052 |
| *WI* | -0.068 | -0.150 | 0.044 | 0.154 | 0.071 | 0.089 | -0.030 | -0.098 |

*Table 1 reports the difference between 2020-Q4 and 2020-Q1 self-reported cancer screening rates for twelve Midwest states. Table 1 also reports the relative change by dividing the Q4-Q1 difference by the Q1 screening rate. All estimates account for sampling probability weights and seasonal differences in response rates.*

**Supplemental Table 2– Change in Self-Reported Difference in Mammograms Rates in 2020 – by Region**

| | Difference | | | Relative Change | | |
|---|---|---|---|---|---|---|
| | Metro | Urban | Rural | Metro | Urban | Rural |
| *IL* | 0.001 | -0.161 | 0.044 | 0.003 | -0.346 | 0.108 |
| *IN* | -0.168 | -0.024 | -0.359 | -0.366 | -0.063 | -0.773 |
| *IA* | -0.098 | -0.012 | 0.152 | -0.265 | -0.045 | 0.547 |
| *KS* | -0.066 | -0.114 | 0.000 | -0.143 | -0.270 | 0.001 |
| *MI* | -0.003 | -0.043 | -0.173 | -0.008 | -0.080 | -0.316 |
| *MN* | -0.022 | 0.057 | 0.008 | -0.059 | 0.144 | 0.021 |
| *MO* | -0.076 | -0.070 | -0.038 | -0.184 | -0.165 | -0.099 |
| *NE* | -0.116 | -0.124 | 0.166 | -0.403 | -0.311 | 0.615 |
| *ND* | -0.067 | -0.061 | -0.007 | -0.164 | -0.151 | -0.020 |
| *OH* | -0.102 | -0.050 | -0.018 | -0.249 | -0.185 | -0.050 |
| *SD* | 0.052 | -0.055 | -0.070 | 0.117 | -0.117 | -0.130 |
| *WI* | -0.057 | -0.223 | 0.056 | -0.123 | -0.452 | 0.162 |

*Table 2 reports the difference between 2020-Q4 and 2020-Q1 self-reported mammogram rates for twelve Midwest states, by metro/urban/rural status of the respondent. Table 2 also reports the relative change by dividing the Q4-Q1 difference by the Q1 screening rate. All estimates account for sampling probability weights and seasonal differences in response rates.*

**Supplemental Table 3– Change in Self-Reported Difference in Pap Smear Rates in 2020 – by Region**

| | Difference | | | Relative Change | | |
|---|---|---|---|---|---|---|
| | **Metro** | **Urban** | **Rural** | **Metro** | **Urban** | **Rural** |
| *IL* | 0.004 | -0.051 | 0.093 | 0.011 | -0.183 | 0.314 |
| *IN* | -0.094 | 0.094 | 0.055 | -0.256 | 0.303 | 0.355 |
| *IA* | -0.017 | -0.108 | 0.073 | -0.054 | -0.353 | 0.255 |
| *KS* | -0.039 | -0.046 | 0.016 | -0.121 | -0.170 | 0.051 |
| *MI* | 0.099 | -0.017 | -0.015 | 0.400 | -0.079 | -0.081 |
| *MN* | -0.010 | -0.027 | 0.001 | -0.032 | -0.092 | 0.006 |
| *MO* | -0.061 | 0.130 | 0.135 | -0.158 | 0.475 | 0.606 |
| *NE* | -0.033 | -0.098 | 0.029 | -0.123 | -0.327 | 0.136 |
| *ND* | -0.022 | 0.042 | 0.055 | -0.058 | 0.180 | 0.231 |
| *OH* | -0.101 | -0.071 | -0.097 | -0.250 | -0.215 | -0.260 |
| *SD* | -0.146 | 0.087 | -0.122 | -0.402 | 0.258 | -0.319 |
| *WI* | 0.014 | 0.089 | 0.129 | 0.044 | 0.461 | 0.517 |

*Table 3 reports the difference between 2020-Q4 and 2020-Q1 self-reported pap smear rates for twelve Midwest states, by metro/urban/rural status of the respondent. Table 3 also reports the relative change by dividing the Q4-Q1 difference by the Q1 screening rate. All estimates account for sampling probability weights and seasonal differences in response rates.*

**Supplemental Table 4– Change in Self-Reported Difference in Blood Stool Test Rates in 2020 – by Region**

| | Difference | | | Relative Change | | |
|---|---|---|---|---|---|---|
| | **Metro** | **Urban** | **Rural** | **Metro** | **Urban** | **Rural** |
| *IL* | 0.033 | 0.004 | -0.029 | 0.042 | 0.004 | -0.037 |
| *IN* | -0.056 | 0.169 | 0.095 | -0.062 | 0.201 | 0.108 |
| *IA* | 0.022 | 0.058 | 0.028 | 0.027 | 0.068 | 0.037 |
| *KS* | 0.051 | -0.051 | -0.067 | 0.070 | -0.063 | -0.084 |
| *MI* | -0.054 | 0.085 | -0.077 | -0.071 | 0.124 | -0.095 |
| *MN* | -0.021 | 0.027 | -0.011 | -0.026 | 0.032 | -0.014 |
| *MO* | -0.035 | -0.018 | -0.042 | -0.042 | -0.022 | -0.052 |
| *NE* | 0.074 | -0.036 | -0.078 | 0.095 | -0.045 | -0.097 |
| *ND* | -0.017 | -0.016 | 0.088 | -0.021 | -0.020 | 0.125 |
| *OH* | 0.058 | -0.087 | -0.075 | 0.074 | -0.104 | -0.091 |
| *SD* | -0.081 | 0.161 | 0.277 | -0.102 | 0.209 | 0.392 |
| *WI* | 0.133 | -0.084 | -0.026 | 0.167 | -0.103 | -0.030 |

*Table 4 reports the difference between 2020-Q4 and 2020-Q1 self-reported blood stool kit test rates for twelve Midwest states, by metro/urban/rural status of the respondent. Table 4 also reports the relative change by dividing the Q4-Q1 difference by the Q1 screening rate. All estimates account for sampling probability weights and seasonal differences in response rates.*

**Supplemental Table 5– Change in Self-Reported Difference in Colon and Sigmoidoscopy Rates in 2020 – by Region**

| | Difference | | | Relative Change | | |
|---|---|---|---|---|---|---|
| | **Metro** | **Urban** | **Rural** | **Metro** | **Urban** | **Rural** |
| *IL* | -0.073 | -0.063 | -0.021 | -0.262 | -0.221 | -0.073 |
| *IN* | 0.008 | -0.323 | 0.032 | 0.027 | -0.700 | 0.293 |
| *IA* | -0.009 | -0.091 | -0.143 | -0.035 | -0.356 | -0.435 |
| *KS* | -0.021 | 0.036 | 0.036 | -0.077 | 0.191 | 0.142 |
| *MI* | -0.085 | -0.120 | -0.052 | -0.256 | -0.350 | -0.165 |
| *MN* | -0.079 | -0.059 | -0.192 | -0.274 | -0.243 | -0.700 |
| *MO* | 0.017 | 0.006 | -0.047 | 0.062 | 0.027 | -0.153 |
| *NE* | -0.141 | 0.157 | -0.121 | -0.401 | 0.721 | -0.395 |
| *ND* | -0.019 | -0.022 | 0.027 | -0.066 | -0.087 | 0.121 |
| *OH* | -0.064 | 0.085 | -0.079 | -0.212 | 0.413 | -0.238 |
| *SD* | 0.066 | -0.031 | -0.082 | 0.180 | -0.103 | -0.233 |
| *WI* | -0.062 | 0.039 | 0.038 | -0.186 | 0.158 | 0.162 |

*Table 5 reports the difference between 2020-Q4 and 2020-Q1 self-reported colon/sigmoidoscopy rates for twelve Midwest states, by metro/urban/rural status of the respondent. Table 5 also reports the relative change by dividing the Q4-Q1 difference by the Q1 screening rate. All estimates account for sampling probability weights and seasonal differences in response rates.*

**Supplemental Table 6 – Change in Self-Reported Blood Stool & Colon/Sigmoidoscopy Rates in 2020 – by Gender**

| | Blood Stool Tests | | | | Colon/Sigmoidoscopies | | | |
|---|---|---|---|---|---|---|---|---|
| | Difference | | Relative Change | | Difference | | Relative Change | |
| | Female | Male | Female | Male | Female | Male | Female | Male |
| *IL* | -0.002 | 0.020 | -0.002 | 0.025 | -0.052 | -0.068 | -0.179 | -0.246 |
| *IN* | -0.076 | 0.020 | -0.083 | 0.022 | 0.104 | -0.118 | 0.382 | -0.402 |
| *IA* | 0.027 | 0.023 | 0.034 | 0.029 | -0.070 | -0.007 | -0.222 | -0.033 |
| *KS* | -0.025 | 0.043 | -0.033 | 0.056 | 0.029 | -0.026 | 0.124 | -0.102 |
| *MI* | -0.078 | -0.010 | -0.102 | -0.013 | -0.088 | -0.068 | -0.240 | -0.233 |
| *MN* | -0.019 | -0.009 | -0.024 | -0.011 | -0.098 | -0.084 | -0.319 | -0.331 |
| *MO* | -0.034 | -0.034 | -0.042 | -0.040 | 0.015 | 0.008 | 0.050 | 0.034 |
| *NE* | -0.038 | 0.034 | -0.047 | 0.043 | -0.024 | -0.036 | -0.073 | -0.136 |
| *ND* | -0.006 | 0.010 | -0.008 | 0.013 | 0.008 | -0.018 | 0.032 | -0.066 |
| *OH* | 0.031 | 0.023 | 0.039 | 0.029 | -0.047 | -0.072 | -0.149 | -0.250 |
| *SD* | 0.034 | 0.095 | 0.044 | 0.123 | 0.082 | -0.043 | 0.204 | -0.147 |
| *WI* | 0.059 | 0.081 | 0.075 | 0.100 | -0.091 | 0.037 | -0.299 | 0.123 |

*Table 6 reports the difference between 2020-Q4 and 2020-Q1 self-reported blood stool test and colon/sigmoidoscopy rates for twelve Midwest states, by self-reported gender of the respondent. Table 6 also reports the relative change by dividing the Q4-Q1 difference by the Q1 screening rate. All estimates account for sampling probability weights and seasonal differences in response rates.*
